# A Machine Learning Framework for Cancer Prognostics: Integrating Temporal and Immune Gene Dynamics via ARIMA-CNN

**DOI:** 10.1101/2024.12.09.24318717

**Authors:** Rui-Bin Lin, Linlin Zhou, Yu-Chun Lin, Yu Yu, Hung-Chih Yang, Chen-Wei Yu

## Abstract

Hepatocellular carcinoma (HCC) poses a significant global health challenge due to its high incidence and mortality rates. Our study investigates the prognostic significance of chemokine (C-C motif) ligand 5 (CCL5) and various immune gene signatures in HCC using an innovative combination of Autoregressive Integrated Moving Average (ARIMA) and Convolutional Neural Network (CNN) models. Time series data were utilized to apply an ARIMA model that captures the temporal dynamics of CCL5 expression. This model’s residual was integrated with immune signature expression data, including lymphocytes and macrophages, to extract features using a CNN model. Our study demonstrates that CNN-extracted features yield a statistically more robust association with patient survival compared to the traditional median split method, which primarily focuses on single-gene analysis. Specifically, CNN-extracted features from CD8 T cells and effector T cells resulted in a hazard ratio (HR) of 0.7324 (*p* = 0.0008) with log-rank *p*-value (0.0131), underscoring their pivotal role in the anti-tumor immune response. This methodology highlights the superior prognostic value obtained through integrated multi-gene analyses, providing deeper insights into tumor-immune interactions than conventional single-gene approaches. Moreover, clustering immune genes based on non-parametric correlations unveiled distinct survival patterns. A cluster comprising B cells, Th2 cells, T cells, and NK cells exhibited a moderate protective effect (HR: 0.8714, *p* = 0.1093) alongside a significant log-rank *p*-value (0.0233). However, the cluster, including granulocytes, Tregs, macrophages, and myeloid-derived suppressor cells, showed no significant survival association, highlighting the intricate immune regulation within the tumor microenvironment. These findings emphasize the necessity of incorporating temporal dynamics and synergistic immune gene interactions for more accurate prognostic evaluations. Our integrated ARIMA-CNN framework represents a significant advancement, leveraging both linear and nonlinear modeling to uncover the dynamic influence of multiple immune genes. This framework holds excellent potential for identifying robust biomarkers and personalizing immunotherapy strategies, ultimately paving the way for innovative cancer management solutions.

## Introduction

Hepatocellular carcinoma (HCC; liver cancer) prevention and cure is the essence of study in the global area of public health. The high incidence and mortality rate of liver cancer necessitates improved prevention, early screening, and treatment strategies to reduce its burden on the public health system. The development of HCC from non-alcoholic fatty liver disease involves the interplay of insulin resistance, inflammatory response to adipose tissue, and several cytokines (*1*). Targeting and addressing potential metabolic disorders, especially those involving inflammation within the tumor immune microenvironment (TIME), may prevent HCC progression and improve patient outcomes (*2*).

Immunotherapy has emerged as a promising avenue for HCC management, with research delving into the TIME. In pathophysiology, the contributions of the TIME, particularly the immune system, including tumor-infiltrating lymphocytes (TILs), cytokine and chemokine networks, play a pivotal role in HCC progression and response to treatment (*3*). Consequently, a comprehensive understanding of immune evasion mechanisms and the development of personalized immunotherapy strategies are not only crucial for achieving optimal treatment outcomes but also play a significant role in extending patients’ overall survival by effectively countering the ability of tumor cells to escape immune surveillance.

Studies have highlighted the potential of time series analysis in improving the management of HCC, including a method that transforms time series data into survival maps. The result has improved dynamic prognosis prediction for patients (*4, 5*). Kaplan-Meier (KM) estimator has been a cornerstone in analyzing survival data, particularly in assessing the impact of genetic expressions on patient outcomes in HCC. However, while KM-estimator analysis is robust for evaluating the levels of single-gene expressions, it may fall short in capturing the complex interplay of multiple genes within the TIME. This limitation can obscure a comprehensive understanding of the intricate immune networks involved in HCC progression and response to treatment.

To address these limitations, advanced time series analysis methods, such as Autoregressive Integrated Moving Average (ARIMA) and ARIMA-Convolutional Neural Network (ARIMA-CNN) models, have emerged as powerful tools for analyzing multiple conditions simultaneously in time series analysis, particularly for complex data sets. ARIMA-CNN leverages ARIMA’s ability to model linear temporal dependencies and CNN’s capability to capture nonlinear patterns and interactions. This hybrid approach provides a more comprehensive and accurate predictive framework, accommodating both linear and nonlinear relationships within the data. The models herein offer a dynamic approach to prognostic prediction, enabling the identification of potential biomarkers that traditional survival analysis techniques could overlook.

Our previous study demonstrated that high chemokine (C-C motif) ligand 5 (CCL5) expression is associated with improved overall survival (OS), disease-free interval, and progression-free interval in HCC, suggesting that CCL5 could serve as a valuable biomarker for patient prognosis (*6*). Here, we propose utilizing the ARIMA-CNN model to analyze the dynamic impact of CCL5 and the other immune genes on HCC patient survival. This innovative application of ARIMA-CNN in the prognostic evaluation of HCC represents a significant advancement in discovering potential biomarkers. It provides a more comprehensive prognostic prediction and personalized treatment strategies. As machine learning continues to evolve, integrating such advanced models into clinical research could revolutionize the management and treatment of HCC, paving the way for more effective and tailored therapeutic interventions.

## Methods

### Acquirement of Immune Gene Profiles from Real-Time Hub Datasets

The mRNA and clinical information of 230 liver tumor patients (n = 230, The Cancer Genome Atlas Liver Hepatocellular Carcinoma) were collected and downloaded from UCSC Xena. The clinical information data includes tumor type, events, overall survival time, and gene expression levels. The expression levels of genes in normal liver tissues were analyzed using data from the Genome Tissue Expression portal. Immune-related genes were screened based on their correlation with immune cell populations in HCC, utilizing data from public medical databases. To categorize gene expression levels, the median and gradient values were used to distinguish between high and low levels of gene expression. \**p* < 0.05 was considered statistically significant.

### Autoregressive Integrated Moving Average-Convolutional Neural Network (ARIMA-CNN) Models

We started by importing essential libraries for our study, including numpy, tensorflow, random, pandas, seaborn, matplotlib, and specific modules from tensorflow.keras and statsmodels.tsa. To ensure the reproducibility of our results, we fixed the random seeds using predefined values (50). The dataset containing CCL5 expression and corresponding survival time data and events was then uploaded. To maintain the integrity of our analysis, we removed any rows with missing values. The ARIMA model was then applied to this time series data with parameters (5,1,0) selected based on preliminary model diagnostics. The residuals of the ARIMA model were computed and stored for further analysis. These residuals represent the differences between the observed values and those predicted by the ARIMA model. The model is defined as *ARIMA* (*p, d, q*), where *p* is the order of the autoregressive part, *d* is the order of differencing, and *q* is the order of the moving average part. For our model, we have *p* = 5, *d* = 1, and *q* = 0. The model is expressed as: Δyt = c+ϕ1Δyt−1+ϕ2Δyt−2+ϕ3Δyt−3+ϕ4Δyt−4+ϕ5Δyt−5+ϵt, where Δyt = yt−yt−1 represents the differenced series, c is a constant, ϕi (for i = 1,…,5) are the coefficients of the autoregressive terms, and ϵt is the white noise error term at time t. The residuals ϵ^t of the ARIMA model are calculated as the difference between the observed values yt and the fitted values y^t: ϵ^t = yt − y^t. These residuals evaluate the model’s performance and ensure that the remaining patterns in the data are random.

To transform the residual data for use in a CNN, we first reshaped the residuals obtained from the ARIMA model into a format suitable for CNN input. Specifically, the residuals were reshaped into a three-dimensional array with shape (len (residuals),1,1), where each dimension has the following interpretation. The first dimension, len (residuals), represents the number of samples in the residual data, with each sample corresponding to a single time step from the ARIMA model. The second dimension denotes the number of features in each residual sample, which is 1 in this case, as the residuals consist of a single numerical value per time step. The third dimension represents the feature depth, with a value of 1 indicating that each feature consists of a single channel, suitable for one-dimensional convolutional operations in the CNN. This reshaping process ensures compatibility with the input requirements of CNN layers, enabling effective extraction of temporal patterns from the residual data.

Next, we included the immune genes as a feature input for the CNN. The immune cell data were extracted from the original dataset and reshaped into a three-dimensional array with shape (features.shape [0], features.shape [1], 1). First dimension features.shape [0], represents the number of samples in the dataset. Each sample corresponds to an observation or instance in the dataset. Second dimension features.shape [1], indicates the number of features for each sample. Since multiple immune genes were included, features.shape [1] allows the representation of multiple genes for each sample. The third dimension represents the feature depth, which is set to 1 because each feature is scalar (a single numerical value), ensuring compatibility with CNN input requirements. A CNN model was then defined using the Sequential API from TensorFlow. The model comprised the following layers:

1. **Conv1D Layer**: A convolutional layer with 32 filters, a kernel size of 1, and Rectified Linear Unit (ReLU) activation function. This layer performs a one-dimensional convolution operation, extracting local patterns from the temporal data.
2. **MaxPooling1D Layer**: A max-pooling layer with a pool size of 1. Since each window contains only one element, the temporal dimension of the data remains unchanged. This layer helps enhance the robustness of the model by reducing sensitivity to minor variations in the data.
3. **Flatten Layer**: A flatten layer to convert the 2D matrix data to a vector, enabling the model to perform regression tasks while retaining all feature values.
4. **Dense Layer**: A fully connected layer with 50 units and ReLU activation function, receiving the flattened vector from the previous Flatten layer and performs deep learning operations and feature extraction.
5. **Output Dense Layer**: A fully connected output layer with 1 unit, enabling one neuron to output one continuous value, and generates the final predictions.

The model was compiled using the Adam optimizer, known for its computational efficiency and adaptive learning rate, and the mean squared error (MSE) loss function, which is suitable for regression tasks. The CNN model was then trained on the input features of immune genes and the reshaped residual data. The training was performed for 50 epochs with a batch size of 10, chosen to balance computational efficiency and convergence stability. After training, the CNN model was used to extract features from the input data. These features represent learned high-level representations of the input data, capturing essential patterns and relationships for subsequent survival analysis. The extracted features were combined with survival time and event data to create a new data frame suitable for survival analysis. Here, the event variable was coded as 1 for death and 0 for follow-up (alive). To ensure data integrity, rows with missing values were removed. This step is essential to maintain the validity of survival analysis, as missing data can bias the results or reduce statistical power.

### Cox Proportional Hazards Model and Kaplan-Meier Estimator

To assess the impact of the CNN-extracted features on survival, we employed the Cox proportional hazards model, a regression method used to explore the relationship between subjects’ survival time and one or more predictor variables under the assumption of proportional hazards. Our study used the lifelines package in Python, a robust library for survival analysis, for this study. We first applied the fit method of the Cox proportional hazards model to the survival data. We employed the Kaplan-Meier estimator further to investigate the impact of the CNN-extracted features on survival. We stratified the data into high and low CNN_features groups based on the median value of CNN_features. Subsequently, Kaplan-Meier survival curves were plotted for these two groups, along with a table displaying the number of subjects at risk at various time points. The complete set of coding scripts, including ARIMA modeling, CNN feature extraction, and clustering algorithms, is hosted on our GitHub repository. All supplementary materials, including detailed code documentation and example datasets, are available upon request.

## Results

### ARIMA Model for CCL5 Expression in HCC

The ARIMA (5, 1, 0) model was fitted to the CCL5 expression time series data, comprising 230 observations. The model demonstrated a log-likelihood of -456.239. The Akaike Information Criterion (AIC) value was 924.479, and the Bayesian Information Criterion (BIC) was 945.081, indicating the model’s relative goodness of fit. The Hannan-Quinn Information Criterion (HQIC) was 932.790, suggesting that the ARIMA (5, 1, 0) model balances model fit and complexity well. This value, along with the AIC and BIC, supports the adequacy of the chosen model for capturing the temporal dynamics of the CCL5 expression data. The autoregressive coefficients (ar.L1 to ar.L4) were all highly significant (*p* < 0.001), suggesting strong temporal dependencies in the CCL5 expression levels. The estimated residual variance (sigma2) was 3.1349 with a standard error of 0.317 (*p* < 0.001), suggesting that the model captures the variability in the CCL5 expression data effectively (Table 1). These results indicate that the residuals are consistent with the model’s assumptions, reinforcing the robustness of the model fit. The Ljung-Box test for autocorrelation in the residuals yielded a Q-statistic of 0.07 with a *p*-value of 0.78, indicating no significant autocorrelation. The Jarque-Bera test for normality of the residuals produced a statistic of 1.1 with a *p*-value of 0.58, suggesting that the residuals follow a normal distribution. The heteroskedasticity test indicated a statistic of 1.21 with a *p*-value of 0.42, suggesting homoskedasticity in the residuals (Table S1). These results collectively indicate that the ARIMA model is an appropriate fit for the CCL5 expression data.

**Table 1.** ARIMA (5, 1, 0) Model Summary.

| Parameter | Coefficient | Std. Error | z-value | $p > \vert z\vert $ | 95% CI |
| --- | --- | --- | --- | --- | --- |
| ar.L1 | -0.7491 | 0.072 | -10.371 | 0 | -0.891 to -0.608 |
| ar.L2 | -0.5649 | 0.083 | -6.829 | 0 | -0.727 to -0.403 |
| ar.L3 | -0.441 | 0.085 | -5.199 | 0 | -0.607 to -0.275 |
| ar.L4 | -0.3706 | 0.082 | -4.512 | 0 | -0.532 to -0.210 |
| ar.L5 | -0.119 | 0.069 | -1.719 | 0.086 | -0.255 to 0.017 |
| sigma2 | 3.1349 | 0.317 | 9.901 | 0 | 2.514 to 3.756 |

### CCL5-ARIMA Model Transforming Residuals and Immune Cell Features for CNN Input

At the beginning of the time series, the model’s fitted values align well with the observed data, reflecting the model’s capability to capture the initial trends and variations. Throughout the middle portion of the time series, the fitted values (red line) continue to track the original data (blue line) with reasonable accuracy (Fig. 1A). The model adapts to the periodic spikes and drops in CCL5 expression, demonstrating its robustness in handling variability. Towards the end of the series, the fitted values show a smooth transition, which may indicate the model’s effort to predict future values based on the learned patterns. The residual plot shows relatively constant variance over time, with no apparent patterns of increasing or decreasing spread (Fig. 1B). This homoscedasticity indicates that the variability in the residuals is stable, which aligns with the assumption of constant variance in the ARIMA model. Overall, the residual analysis supports the adequacy of the ARIMA model for the CCL5 expression data. The autocorrelation and partial autocorrelation analyses, as illustrated in Fig. S1A and B, support the robustness and reliability of the ARIMA model. The residuals lack significant and partial autocorrelations, confirming that the model’s assumptions are satisfied and adequately capture the temporal dynamics of CCL5 expression. These findings, combined with the statistical significance of the model parameters and the excellent fit to the original data, underscore the robustness and reliability of the ARIMA model in capturing the temporal dynamics of CCL5 expression.

**Figure 1.**
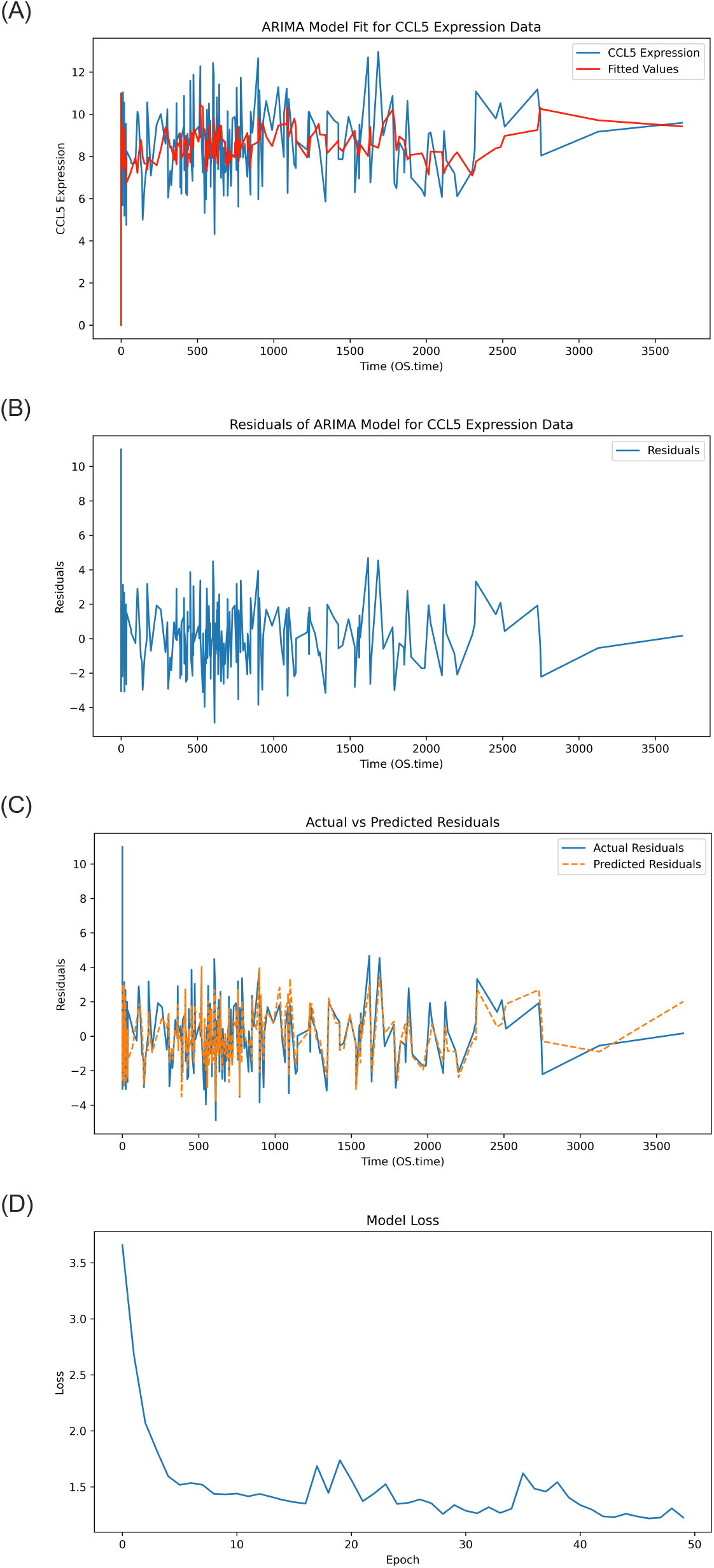
ARIMA model fit for CCL5 expression and the immune cell expressions for CNN. (A) The blue line represents the original CCL5 expression data, while the red line indicates the fitted values generated by the ARIMA (5, 1, 0) model. (B) The residuals over time are the differences between the observed CCL5 expression levels predicted by the ARIMA model. (C) The residuals from the ARIMA model were reshaped and plotted to visualize their suitability as input data for a CNN. The Blue solid line is the actual residuals obtained from the ARIMA model applied to the CCL5 data, and the orange dashed line is the predicted residuals generated by the CNN model using multiple gene features. (D) The Y-axis represents the training loss, which measures the model’s prediction error on the training data, and the X-axis represents the number of training epochs. OS.time as overall survival time (days).

While ARIMA models handle linear relationships proficiently, many genetic expression patterns are inherently nonlinear, particularly in TIME. The model can learn and capture the nonlinear relationships that ARIMA models might miss by inputting the residuals into a CNN. Therefore, transforming residuals for CNN input may provide several advantages, including capturing nonlinear relationships, complementing or augmenting the ARIMA model, and improving prediction accuracy. Immune gene interactions can be complex and interdependent. Multiple gene features allow the model to capture these interactions, which is critical for the accuracy of survival prediction. The comparison of actual residuals and predicted residuals was therefore analyzed. To verify whether there is a potential problem of overfitting, we applied all selected immune genes including CD8 T cells, granzyme B (GZMB), perforin 1 (PRF1), T helper type 1 cells (Th1), B cells, T cells, T helper type 2 cells (Th2), natural killer (NK) cells, granulocytes, regulatory T cells (Tregs), macrophages and myeloid-derived suppressor cells (MDSCs) to the CCL5-ARIMA model for a cross-validation method to address the issue. The average training loss is 1.182, and the average validation loss is 1.346. From the results, we calculated the loss ratio of 0.139 (13.9%), which is relatively small when considering our medium-sized dataset, suggesting that the validation loss remains reasonably close to the training loss to some extent (Fig. S1C). The green dashed line represents the validation loss curve, while the blue dashed line represents the training loss curves. The two curves show the decreasing trend over the epoch, as observed from the yellow solid line, indicating no significant sign of overfitting. Combining the loss ratio and average training and validation loss curves, we conclude that no significant effect of overfitting was observed in our CNN model. Next, the ARIMA model residuals were reshaped and plotted to visualize the suitability for a CNN (Fig. 1C). The blue solid line represents the actual residuals obtained by applying the ARIMA model to the CCL5 data. The orange dashed line represents the predicted residuals generated by the CNN model using all selected genes. The predicted residuals closely follow the actual residuals at most time points, indicating that the CNN model has successfully captured the residual patterns to a considerable extent. The CNN model’s training loss over epochs was trained using the transformed ARIMA residuals and the other immune gene expression data as inputs. The training loss decreases rapidly during the initial epochs, indicating that the model quickly learns valuable features from the data. As the number of epochs increases, the loss gradually stabilizes, suggesting that the model’s learning process has reached a steady state. The final loss value is low and stable, demonstrating that the model fits well with the training data (Fig. 1D). The CNN model is subsequently applied to extract features from the input data, comprising the transformed ARIMA residuals and immune cell expression profiles. These extracted features are designed to capture complex patterns and non-linear interactions, providing insights that are predictive of survival outcomes.

### ARIMA-CNN-Driven Insights into Immune Gene Interactions and Prognostic Survival in HCC

To evaluate the ARIMA-CNN model’s predictive power and stratification capability, Kaplan-Meier survival curves were analyzed and stratified based on the median expression levels of CCL5, the expression levels of other immune cells, and the CNN-extracted features derived from the CCL5-ARIMA model residuals. This analysis highlights the comparative performance of traditional median-based stratification versus CNN-enhanced feature extraction in predicting survival outcomes. The median split analysis of single genes revealed significant survival associations for specific immune gene signatures, including B cells, Th1 cells, CD8 T cells, and Tregs. These findings highlight the potential prognostic value of individual immune gene expressions when stratified by their median expression levels (Table S2). In contrast, the CNN-based feature extraction method, leveraging CCL5-ARIMA residuals, revealed a consistent trend of reduced hazard ratios (HR), indicating a stronger protective effect compared to the median split analysis. For instance, the HR for CD8 T cells decreased from 0.8037 to 0.7654, and for Th1 cells from 0.7485 to 0.7720 (Table S3). These reduced hazard ratios highlight the ability of the CNN model to effectively capture complex, non-linear patterns in the data, which are not apparent in traditional stratification methods. Furthermore, the CNN-extracted features demonstrated that CCL5 and its associated immune gene signatures, such as CD8 T cells and Th1 cells, synergistically influence patient outcomes by providing a more nuanced representation of survival-related features. Herein, this finding underscores the model’s ability to integrate interactions between multiple immune gene expressions, offering a comprehensive understanding of their prognostic value and surpassing the predictive capability of traditional single-gene median split approaches.

An intriguing consistency in log-rank *p*-values was observed between the median split and CNN-extracted feature analyses. This phenomenon suggests that while the CNN model captures more complex patterns and interactions between genes, it still retains critical characteristics of the raw gene expression data. As a result, the stratification of survival times based on CNN features aligns closely with the original median split analysis, particularly for genes with strong linear survival signals. Building on this observation, we investigate the linear correlation between CCL5 and other immune genes to better understand their collective impact on survival outcomes. The moderate to strong positive correlations between CCL5 and other immune genes suggest that linear relationships exist (Fig. S2). To further refine our approach, we propose clustering the immune genes based on their Spearman rho correlations and then using these clusters for CCL5-ARIMA and CNN-based survival analysis. Using hierarchical clustering, we group the genes into three distinct clusters based on their correlation patterns (Table 2). We perform ARIMA modeling on CCL5 expression data for each cluster to extract residuals. These residuals and gene expressions in the clusters are then used as input features for a CNN model. Our results show that an HR of 0.8714 with a log-rank *p*-value of 0.0233 for immune cell panel 2 suggests a protective effect; however, the Cox *p*-value of 0.1093 indicates borderline statistical significance, which limits the robustness of this finding. Furthermore, the non-significant results for panel 3 (Cox *p*-value of 0.4276 and log-rank *p*-value of 0.7894) suggest that when analyzed in isolation, these genes have limited or no prognostic value in relation to CCL5 and patient survival (Table 2). In contrast, immune cell panel 1 demonstrates a stronger association with survival outcomes, with a Cox *p*-value of 0.0008 and a log-rank *p*-value of 0.0131. This result, coupled with the reduced HR (0.7324, 95% CI: 0.6101 - 0.8793), indicates a more robust prognostic value than individual genes. Notably, genes in panel 1 show significant correlations with CCL5-ARIMA residuals, suggesting that these genes may interact with the temporal dynamics of CCL5 expression to influence patient survival collectively (Table 2). These findings indicate that while specific immune gene panels exhibit limited prognostic utility, panel 1 represents a more reliable indicator of survival due to its more decisive statistical significance, protective effect, and integration with the temporal dynamics captured by the CCL5-ARIMA model. The three panels’ Kaplan-Meier Estimator and risk counts were also performed and aligned with the statistical results (Fig. 2). Taken together, these results underscore the complexity of immune regulation within the TIME and highlight the importance of evaluating gene interactions rather than isolated effects to fully understand their role in survival outcomes.

**Table 2.**
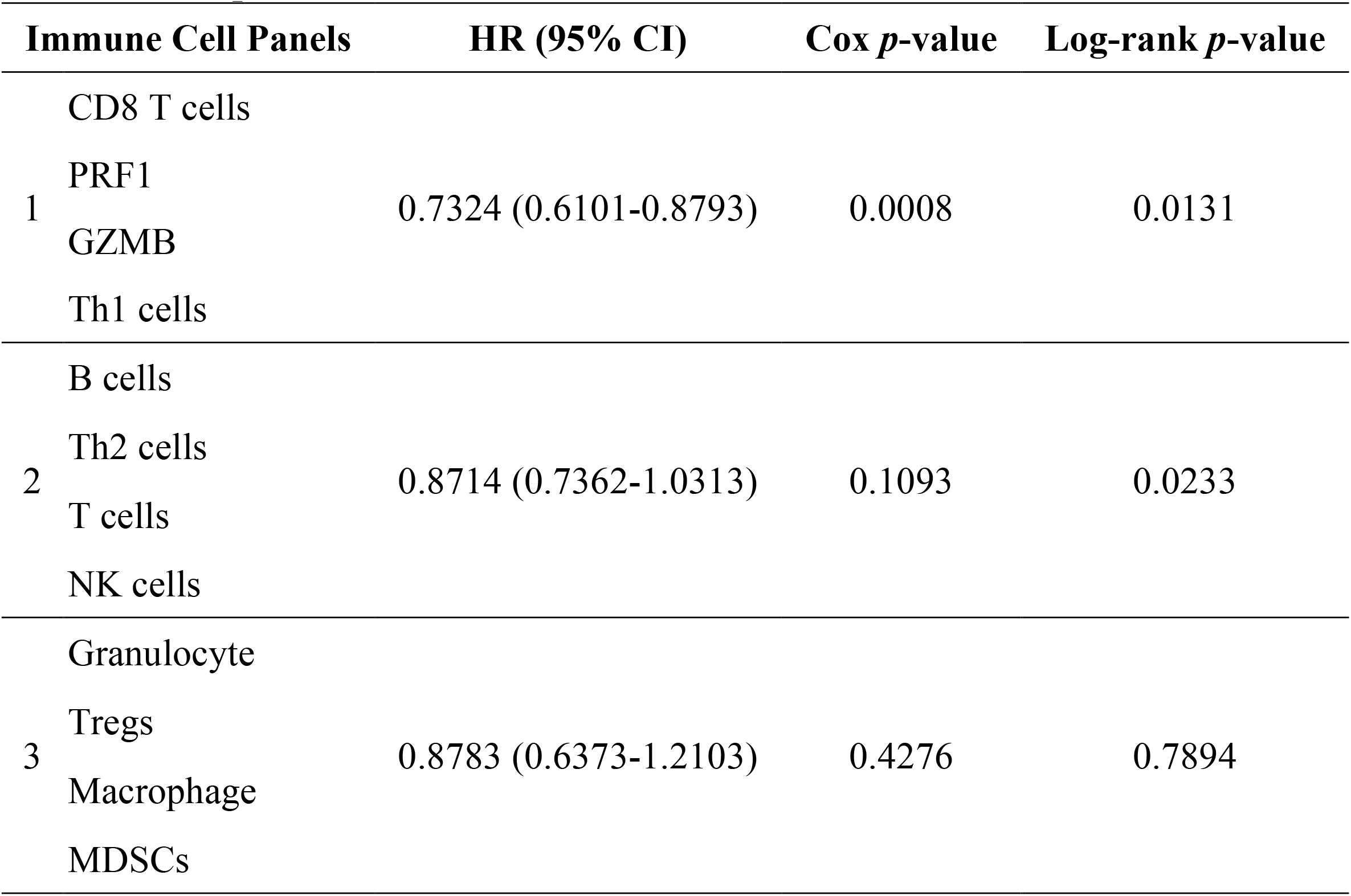
Multiple Genes of CCL5-ARIMA CNN-Extracted Features.

**Figure 2.**
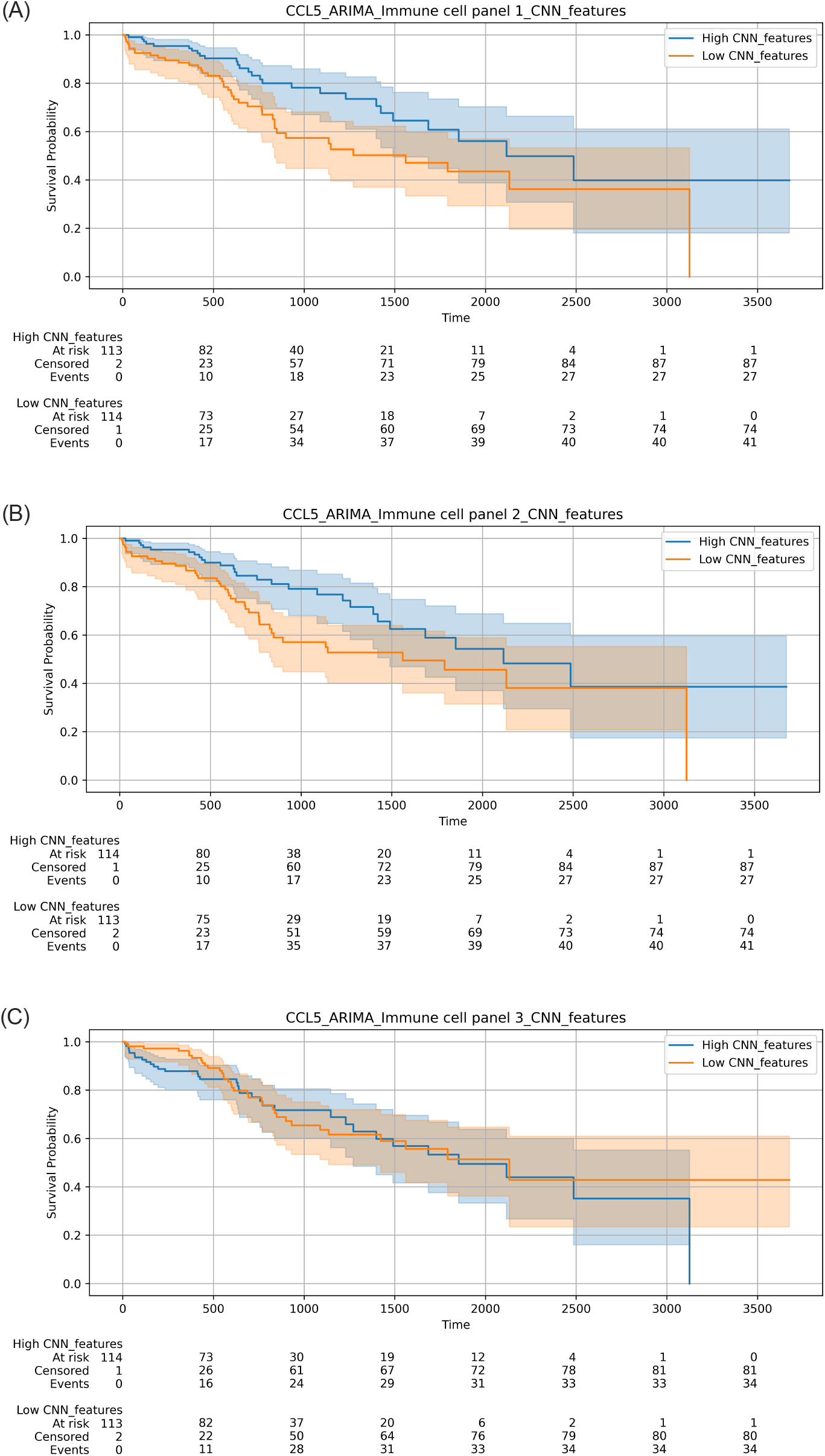
Kaplan-Meier Survival Curves for CCL5-ARIMA CNN-Extracted Immune Gene Clusters. (A) The overall survival of HCC patients was stratified by high and low CNN-extracted features from immune gene signatures, specifically CD8 T cells, PRF1, GZMB, and Th1 cells. Patients with high CNN features (blue line) demonstrated significantly better survival compared to those with low CNN features (orange line), with a log-rank *p*-value of 0.0131. (B) HCC patients were stratified by high and low CNN-extracted features from the cluster of B cells, Th2 cells, T cells, and NK cells. The significant log-rank *p*-value of 0.0233 suggests notable differences in survival between the groups. (C) HCC patients were stratified by high and low CNN-extracted features from granulocytes, Tregs, macrophages, and MDSCs. No significant difference was observed in survival between the groups. The risk counts were present in the figures.

## Discussion

Gene expression is crucial in understanding various diseases’ biological mechanisms, including cancer. The combination of time series models and machine learning techniques can provide valuable insights into the predictive power of gene expression profiles. Our study investigated the prognostic significance of CCL5 time series expression and its association with various immune gene signatures in cancer patient survival analysis. Our approach integrated ARIMA modeling, CNN-extracted features, and survival analysis to understand these factors’ relationships comprehensively. Our initial step involved constructing an ARIMA model for CCL5 time series expression to account for temporal dependencies and extract residuals. The ARIMA model revealed significant temporal patterns in CCL5 expression, with the residuals representing the mysterious variance. These residuals were subsequently used as inputs for CNN models alongside immune gene expressions to predict survival outcomes.

The ARIMA model, a crucial tool in our research, effectively captured the dynamic temporal trends in CCL5 expression, thereby highlighting its significant role in understanding HCC progression. We then assessed the prognostic value of individual immune genes by integrating their expressions with CCL5-ARIMA residuals and extracting features using CNNs. Notably, several immune genes, including CD8 T cells, Th1 cells, and B cells, exhibited significant associations with survival. Other immune signatures, such as PRF1 and GZMB, showed trends toward significance, indicating their potential importance in the immune response against HCC. However, the granulocyte and macrophage did not show significant associations, suggesting that their roles might be more context-dependent or complex. From the linear analysis, CCL5 showed moderate to strong positive correlations with several immune genes, particularly CD8 T cells. These correlations suggested that genes with similar expression patterns might be co-regulated or involved in related biological pathways, justifying the subsequent clustering approach.

We herein grouped immune genes into clusters and conducted survival analysis using CNN-extracted features from these clusters. This approach aimed to capture the combined effects of biologically related genes. The ARIMA model of CCL5 accounts for its temporal variations, and the residuals used in the CNN capture the mysterious variance after accounting for these variations. By integrating these residuals with the expression levels of panel 1 immune genes, the model effectively encapsulates the dynamic interplay between CCL5-driven processes and the immune response mediated by these immune signatures, including CD8 T cells, effect T cells (PRF1^+^/GZMB^+^), and Th1 cells. This interplay is crucial for enhancing our understanding of their combined prognostic value. The moderate significance of panel 2 genes, when combined with CCL5-ARIMA residuals, suggests that while these immune cells, including B cells, Th2 cells, T cells, and NK cells, do play a role in the immune response, their temporal dynamics and interaction with CCL5 are less pronounced than those in panel 1. Interestingly, the lack of statistical significance in panel 3, despite integrating CCL5-ARIMA residuals and their strong linear correlation with CCL5 expression, underscores the complexity of immune regulation within the TIME. While the ARIMA residuals capture the temporal dynamics of CCL5 expression, the heterogeneity among the included immune cell types, such as granulocytes, Tregs, macrophages, and MDSCs, may dilute their collective impact on survival outcomes. These cell types likely exert diverse and context-dependent effects on tumor progression and immune modulation, which could mask the individual contributions of CCL5 when analyzed together.

The ARIMA model of CCL5 captures its temporal expression patterns, which is crucial for understanding how CCL5 influences the TIME over time. Using the residuals from this model, we ensure that the subsequent analyses focus on the interactions between CCL5 and immune genes beyond their individual expressions. The significant findings in cytotoxic T lymphocytes (CD8 T cells) demonstrate that genes involved in the cytotoxic immune response work synergistically with CCL5 dynamics to enhance survival. This synergy indicates a robust anti-tumor immune environment driven by both the temporal expression of CCL5 and the active roles of cytotoxic immune cells. To further demonstrate the advantages of ARIMA-CNN modeling in integrating multiple genes for survival analysis, we compared its performance with traditional Kaplan-Meier curve stratification based on the expression levels of CCL5 and CD8 T cells. Four survival groups were generated using the Kaplan-Meier approach based on high and low CCL5 and CD8 T cell expression levels. The Kaplan-Meier analysis demonstrated statistical significance in only two survival curve groups: the red curve (high CCL5 & high CD8 T cells) compared to the green curve (high CCL5 & low CD8 T cells) with a Cox *p* = 0.007 and log-rank *p* = 0.004, and the red curve compared to the blue curve (low CCL5 & low CD8 T cells) with a Cox *p* = 0.014 and log-rank *p* = 0.012 (Fig. S3A). However, the remaining groups did not show significant differences, reflecting the inherent limitations of traditional stratification methods in capturing the complex interplay between genes. Furthermore, Kaplan-Meier methods rely heavily on pre-defined groupings (e.g., high/low expression levels) and require multiple rounds of stratification and comparisons, making the process both labor-intensive and less flexible when dealing with intricate gene interactions. This complexity increases the risk of overlooking subtle but biologically meaningful patterns in the data. In contrast, the ARIMA-CNN approach simplifies the analytical workflow by automatically integrating temporal patterns (via ARIMA) and extracting non-linear interactions (via CNN). ARIMA-CNN provides a more comprehensive and interpretable assessment of survival-related features by directly quantifying the interplay between CCL5’s temporal expression dynamics and CD8 T cells (Fig. S3B), offering more profound insights into their collective impact on patient survival. Our findings demonstrate that ARIMA-CNN not only reduces the manual effort required for group-based stratification but also provides deeper insights into gene interactions, showing its superiority in both efficiency and prognostic power.

These results underscore the transformative potential of ARIMA-CNN as a novel approach to survival analysis, particularly for exploring the dynamic interplay of multiple genes over time. Unlike traditional methods like Kaplan-Meier curves, which are limited to predefined categorical groupings, ARIMA-CNN integrates temporal dynamics and multi-gene interactions, offering deeper biological insights and significantly improving prognostic modeling. Our findings demonstrate that while specific immune genes, such as CD8 T cells, effector T cells, Th1 cells, and B cells, exhibit strong protective effects and critical roles in the immune response against tumors, the combined effects of other immune cells, such as granulocytes and MDSCs, are less pronounced (*7*). By leveraging ARIMA, the temporal dynamics of CCL5 expression were effectively captured, and CNN-extracted features further revealed significant associations between immune gene clusters and patient survival (Fig. 3) (*8*).

**Figure 3.**
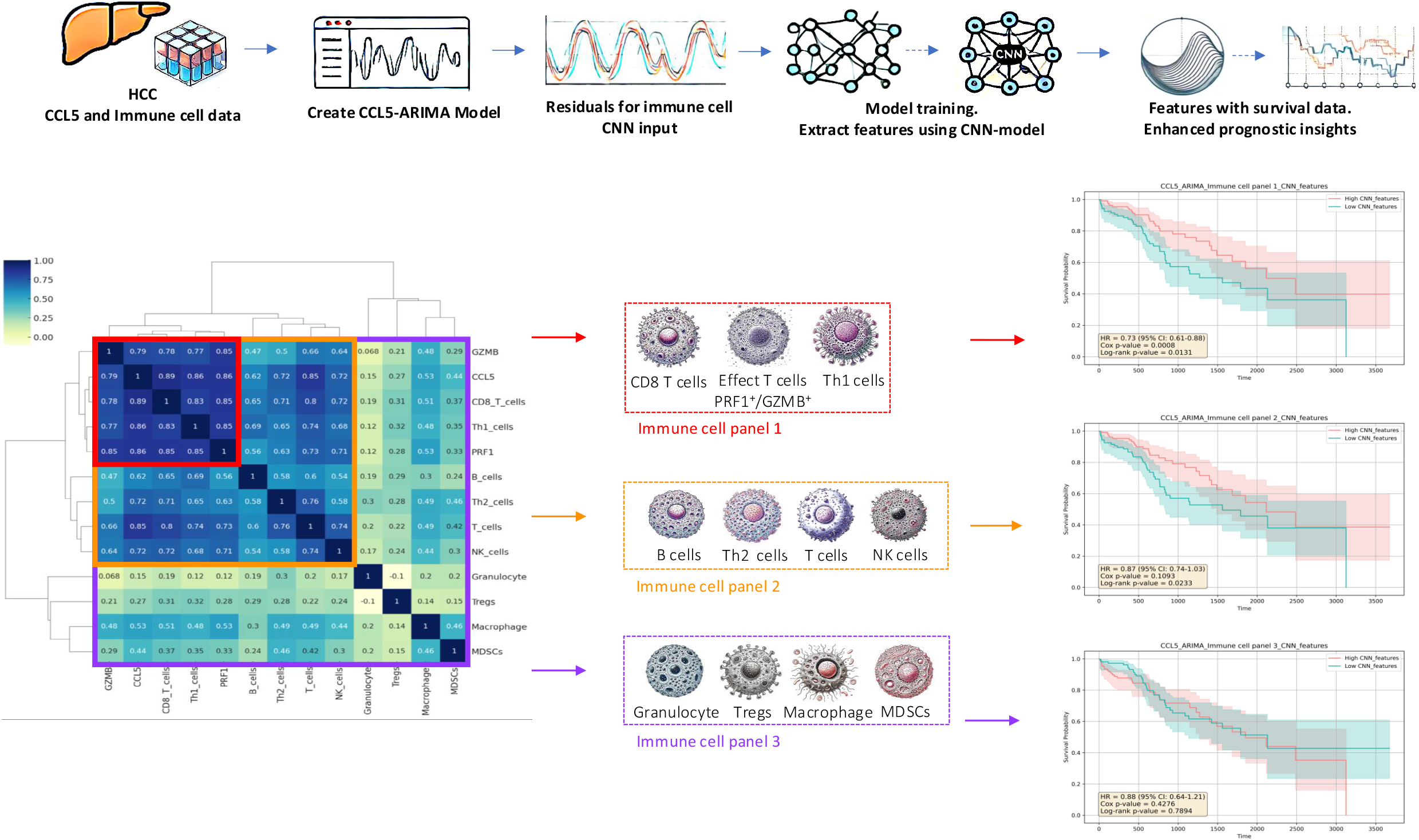
The pipeline of the ARIMA-CNN model in HCC immune signature analysis. The process begins with creating the CCL5-ARIMA model to handle the temporal expression data of CCL5 in HCC patients. The residuals from the ARIMA model are used as inputs for training a CNN, which also integrates immune cell gene expression data. The CNN extracts features that are subsequently combined with survival data to enhance prognostic insights. The immune genes are clustered into three panels based on Spearman correlation: Panel 1 as red (CD8 T cells, PRF1, GZMB, Th1 cells), Panel 2 as orange (B cells, Th2 cells, T cells, NK cells), and Panel 3 as violet (Granulocyte, Tregs, Macrophage, MDSCs). Each panel’s prognostic significance is analyzed, with Panel 1 showing a highly significant protective effect on survival, Panel 2 demonstrating a moderate protective effect, and Panel 3 does not show significant associations. This approach highlights the impact of immune gene interaction within CCL5 dynamics on patient survival.

Notably, ARIMA-CNN provides a powerful framework for analyzing multiple genes simultaneously, particularly in cases where the prognostic significance of these genes is not well established. This approach enables researchers to identify gene expression patterns and interactions that are significantly associated with survival outcomes, providing a basis for future investigations into the role of these gene clusters in cancer immunity. Moreover, it paves the way for targeted therapeutic interventions, focusing on specific immune gene clusters to enhance the effectiveness of cancer immunotherapy. We herein highly recommend leveraging machine learning approaches such as ARIMA-CNN to analyze temporal and hybrid gene features. This method not only delivers statistically significant prognostic information but also uncovers critical biological insights, offering a foundation for advancing precision oncology and guiding therapeutic strategies.

## Data Availability

All data produced in the present study are available upon reasonable request to the authors.

