## Supplementary material for "A Machine Learning Framework for Cancer Prognostics: Integrating Temporal and Immune Gene Dynamics via ARIMA-CNN": Fig. S1

### Supplementary Figures

(A)

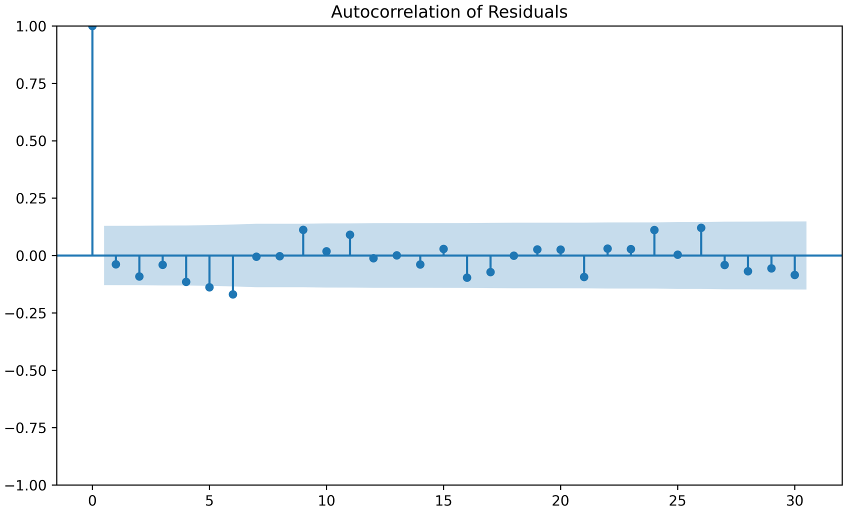


(B)

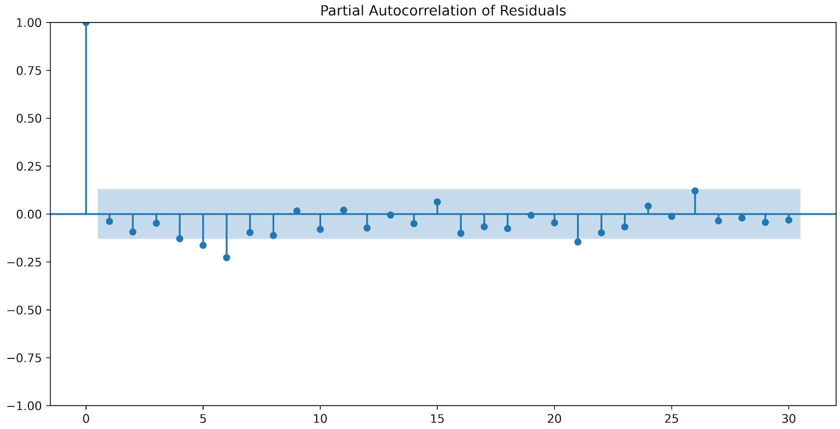


(C)

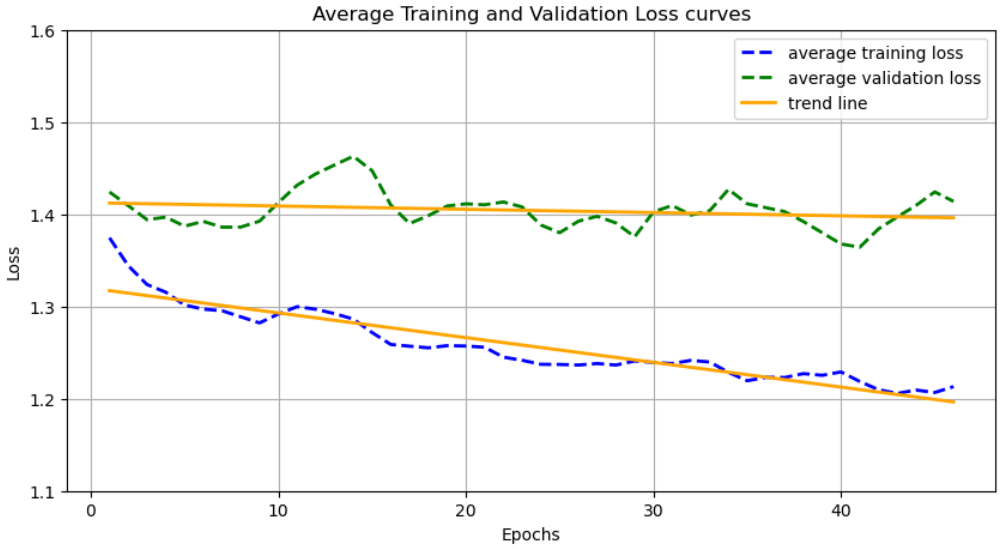

**Figure S1. Residuals of ARIMA Model for CCL5 Expression Data.**

(A) The autocorrelation function (ACF) plot shows that most of the autocorrelation coefficients are within the 95% confidence interval bounds, suggesting that there is no significant autocorrelation present in the residuals. This observation is consistent with the results of the Ljung-Box test, which did not indicate significant autocorrelation (Prob(Q) = 0.78). (B) Similar to ACF, the partial autocorrelation function (PACF) plot shows that the partial autocorrelation coefficients are within the 95% confidence interval bounds, further supporting the absence of significant partial autocorrelations. This indicates that any remaining structure in the residuals is negligible and does not affect the model's adequacy. (C) Cross-validation is applied to verify the potential problem of overfitting. The loss ratio of 0.139 (13.9%) is relatively small, especially considering our dataset is not particularly large, suggesting that the validation loss remains reasonably close to the training loss. The two-loss curves show a decreasing trend over the epoch, as observed from the yellow solid line. From the results, we conclude that overfitting has a non-significant effect on our CNN model.
