## Supplementary material for "A Machine Learning Framework for Cancer Prognostics: Integrating Temporal and Immune Gene Dynamics via ARIMA-CNN": Fig. S2

(A)

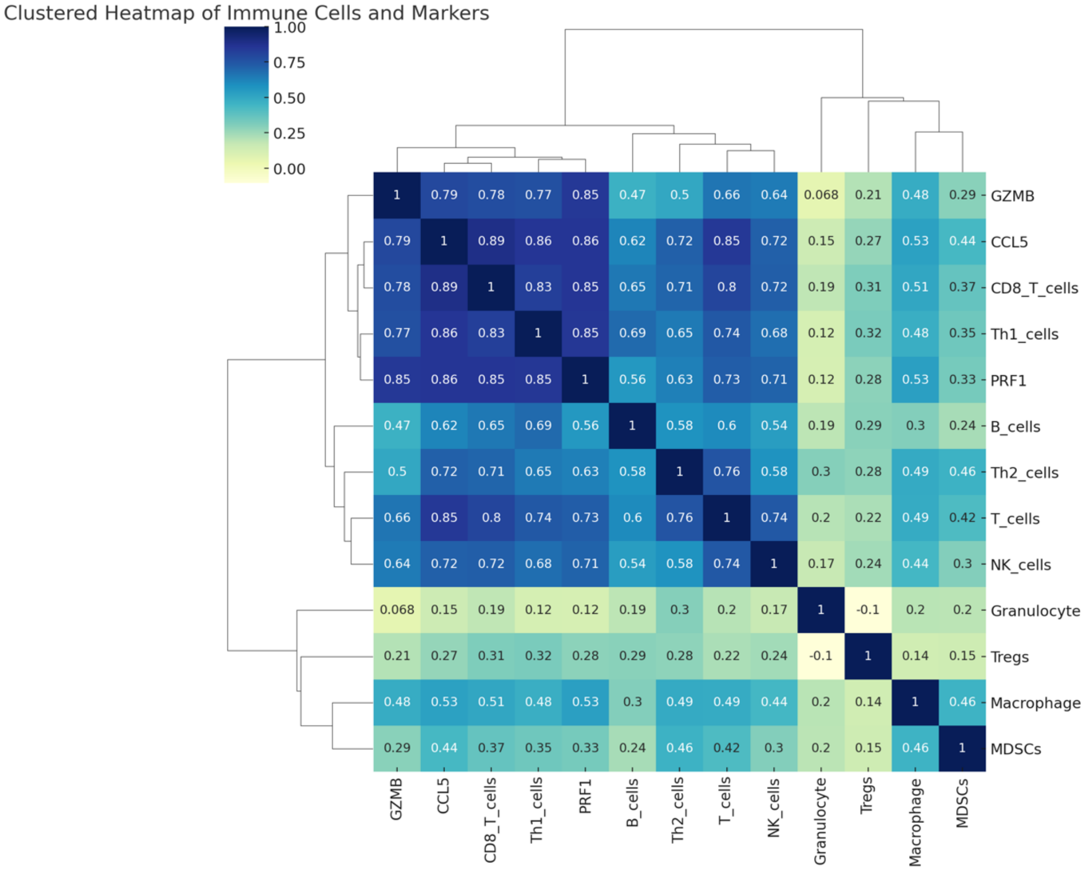
Clustered Heatmap of Immune Cells and Markers
1.00
0.75
0.50
0.25
0.00
GZMB
CCL5
CD8_T_cells
Th1_cells
PRF1
B_cells
Th2_cells
T_cells
NK_cells
Granulocyte
Tregs
Macrophage
MDSCs

(B)

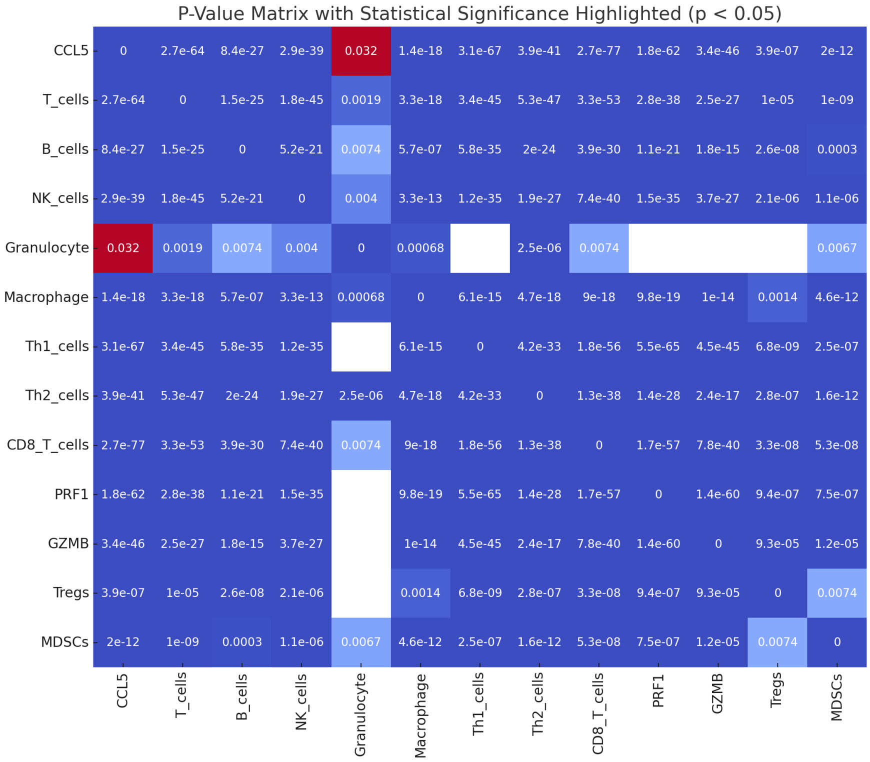
P-Value Matrix with Statistical Significance Highlighted (p < 0.05)
CCL5
T_cells
B_cells
NK_cells
Granulocyte
Macrophage
Th1_cells
Th2_cells
CD8_T_cells
PRF1
GZMB
Tregs
MDSCs

**Figure S2. The non-parametric correlation analysis between CCL5 expression and various immune cells.**

(A) Non-parametric Spearman rho between CCL5 and the selected immune genes are calculated and analyzed to determine the strength and direction of their linear relationships. (B) The correlation of the *p*-value matrix with incorporated in the figure.
