## Supplementary material for "A Machine Learning Framework for Cancer Prognostics: Integrating Temporal and Immune Gene Dynamics via ARIMA-CNN": Fig. S3

(A)

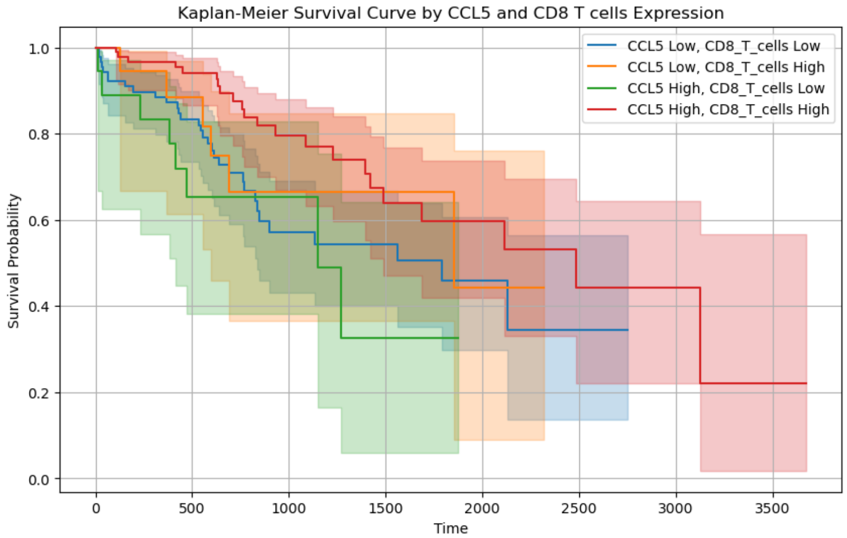


(B)

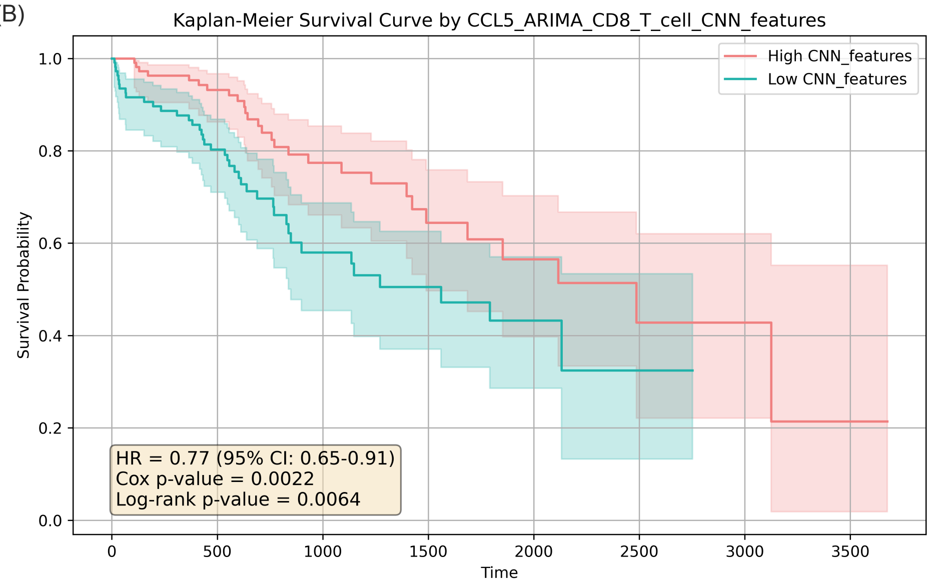


**Figure S3. Kaplan-Meier Survival Curve and Cox Regression Analysis for CCL5-ARIMA CD8 T cells CNN-Extracted Features.**

**(A)** The result of the Kaplan-Meier curves graph appears to be more complicated when compared to CCL5 and CD8 T cells, with only two survival curve groups showing significant differences. (B) The ARIMA modeling on CCL5 expression data for CD8 T cells to extract residuals was used as input features for the CNN model alongside gene expression.
