## Supplementary material for "A Machine Learning Framework for Cancer Prognostics: Integrating Temporal and Immune Gene Dynamics via ARIMA-CNN": Table S1

**Supplementary Tables**

**Table S1. Diagnostic Tests**

| Test | Statistic | Prob. |
|---|---|---|
| Ljung-Box (L1) (Q) | 0.07 | 0.78 |
| Jarque-Bera (JB) | 1.1 | 0.58 |
| Heteroskedasticity (H) | 1.21 | 0.42 |
