## Supplementary material for "A Machine Learning Framework for Cancer Prognostics: Integrating Temporal and Immune Gene Dynamics via ARIMA-CNN": Table S2

**Table S2. Median Split Analysis of Single Gene.**

| Immune signatures | HR | 95% CI | Cox *p*-value | Log-rank *p*-value |
|---|---|---|---|---|
| CCL5 | 0.8638 | 0.7428-1.0045 | 0.0572 | 0.0777 |
| T cells | 0.9366 | 0.8320-1.0544 | 0.2785 | 0.1115 |
| B cells | 0.8581 | 0.7673-0.9595 | 0.0073 | 0.0087 |
| NK cells | 0.9486 | 0.8175-1.1007 | 0.4869 | 0.7471 |
| Granulocyte | 1.0565 | 0.9130-1.2225 | 0.4605 | 0.5039 |
| Macrophage | 1.0082 | 0.8026-1.2663 | 0.9442 | 0.2526 |
| Th1 cells | 0.7485 | 0.6226-0.8998 | 0.0020 | 0.0045 |
| Th2 cells | 0.9907 | 0.8549-1.1481 | 0.9011 | 0.5972 |
| CD8 T cells | 0.8037 | 0.7002-0.9224 | 0.0019 | 0.0064 |
| PRF1 | 0.8473 | 0.7050-1.0182 | 0.0770 | 0.2426 |
| GZMB | 0.8601 | 0.7412-0.9980 | 0.0471 | 0.2017 |
| Tregs | 0.8727 | 0.7774-0.9796 | 0.0210 | 0.0343 |
| MDSCs | 1.0723 | 0.9140-1.2580 | 0.3915 | 0.8359 |
