## Supplementary material for "A Machine Learning Framework for Cancer Prognostics: Integrating Temporal and Immune Gene Dynamics via ARIMA-CNN": Table S3

**Table S3. CCL5-ARIMA-CNN Extracted Features of Single Gene.**

| Immune signatures | HR | 95% CI | Cox *p*-value | Log-rank *p*-value |
|---|---|---|---|---|
| T cells | 0.9262 | 0.7931-1.0817 | 0.3328 | 0.1115 |
| B cells | 0.7322 | 0.5835-0.9189 | 0.0072 | 0.0087 |
| NK cells | 0.9278 | 0.7522-1.1445 | 0.4842 | 0.7471 |
| Granulocyte | 2.9884 | 0.1664-53.6804 | 0.4576 | 0.5039 |
| Macrophage | 1.1190 | 0.0480-26.1099 | 0.9442 | 0.2526 |
| Th1 cells | 0.7720 | 0.6462-0.9222 | 0.0043 | 0.0045 |
| Th2 cells | 0.9704 | 0.7959-1.1831 | 0.7662 | 0.5972 |
| CD8 T cells | 0.7654 | 0.6450-0.9082 | 0.0022 | 0.0064 |
| PRF1 | 0.8568 | 0.7165-1.0247 | 0.0905 | 0.2426 |
| GZMB | 0.8278 | 0.6810-1.0064 | 0.0579 | 0.2017 |
| Tregs | 0.5354 | 0.3088-0.9283 | 0.0261 | 0.0343 |
| MDSCs | 1.1618 | 0.7977-1.6920 | 0.4343 | 0.8359 |
